# Sleep and long COVID: Preexisting sleep issues and the risk of PASC in a large general population using 3 different model definitions

**DOI:** 10.1101/2024.06.20.24309263

**Authors:** Stuart F. Quan, Matthew D. Weaver, Mark É. Czeisler, Laura K. Barger, Lauren A. Booker, Mark E. Howard, Melinda L. Jackson, Rashon I. Lane, Christine F. McDonald, Anna Ridgers, Rebecca Robbins, Prerna Varma, Joshua F. Wiley, Shantha M.W. Rajaratnam, Charles A. Czeisler

## Abstract

**Study Objectives:** Insomnia, poor sleep quality and extremes of sleep duration are associated with COVID-19 infection. This study assessed whether these factors are related to Post-Acute Sequelae of SARS-CoV-2 infection (PASC).

**Methods:** Cross-sectional survey of a general population of 24,803 U.S. adults to determine the association of insomnia, poor sleep quality and sleep duration with PASC.

**Results:** Prevalence rates of PASC among previously COVID-19 infected participants for three definitions of PASC were COPE (21.9%), NICE (38.9%) and RECOVER PASC Score (15.3%). PASC was associated with insomnia in all 3 models in fully adjusted models with adjusted odds ratios (aORs) and 95% confidence intervals (CI) ranging from 1.30 (95% CI: 1.11-1.52, p≤0.05, PASC Score) to 1.52 (95% CI: 1.34-1.71, p≤0.001, (NICE). Poor sleep quality was related to PASC in all models with aORs ranging from 1.77 (95% CI: 1.60-1.97, p≤0.001, NICE) to 2.00 (95% CI: 1.77-2.26, p≤0.001, COPE). Sleep <6 hours was associated with PASC with aORs between 1.59 (95% CI: 1.40-1.80, p≤0.001, PASC Score) to 1.70 (95% CI: 1.53-1.89, p≤0.001, COPE). Sleep ≥ 9 hours was not associated with PASC in any model. Although vaccination with COVID-19 booster decreased the likelihood of developing PASC, it did not attenuate associations between insomnia, poor sleep quality and short sleep duration with PASC in any of the models.

**Conclusions:** Insomnia, poor sleep quality and short sleep duration are potential risk factors for PASC. Interventions to improve sleep may decrease the development of PASC.

**Brief Summary:** *Current Knowledge/Study Rationale:* Insomnia, poor sleep quality, and extremes of sleep duration have been associated with a higher likelihood of COVID-19 infection. However, evidence implicating an association with the development of Post-Acute Sequelae of SARS-CoV-2 infection (PASC) is scant.

*Study Impact:* Results indicate that insomnia, poor sleep quality and sleep duration ≤6 hours are associated with an increase in the prevalence of PASC among persons who have previously had a COVID-19 infection. The findings provide support for employing interventions to improve sleep as a means to decrease the development of PASC.

## Introduction

As the COVID-19 pandemic has evolved into global endemic status, the emergence of persistent and/or relapsing-remitting physical, cognitive and mental health symptoms commonly known as “long COVID” or more formally as Post-Acute Sequelae of SARS-CoV-2 infection (PASC) has become an increasingly important public health concern. ^1–4^ Risk factors for the development of PASC include greater severity of initial COVID-19 illness, multiple infections, female sex and preexisting health conditions.^2,5^ Recently, obstructive sleep apnea (OSA) has been demonstrated in two large studies to be associated with the development of PASC.^6,7^ Although poor sleep quality and short sleep duration are associated with an increased risk of acute COVID-19 infection,^8,9^ it is uncertain whether they also are related to a higher likelihood of PASC.

Estimates of the prevalence of PASC are imprecise, ranging from 7.5 to 41%.^10^ In part, this imprecision is related to the lack of a generally accepted definition of this syndrome. Several definitions have been proposed that include the use of various combinations of duration of time post infection, number of symptoms and a scoring system that weights some symptoms more than others.^6,11–14^ However, there are few data that have prospectively tested or compared definitions to each other, control groups and long-term outcomes.

In this study, we aimed to determine in a large general population cohort whether preexisting insomnia, poor sleep quality, and short or long sleep duration are associated with the development of PASC using 3 different model definitions of the syndrome. In addition, we compared the prevalence of PASC among these models.

## Methods

### Study Design and Participants

From March 10 to October 15, 2022, the COPE Initiative^15^ administered five successive waves of surveys focused on accumulating data on the prevalence and sequelae of COVID-19 infection in the United States (U.S.). Dates of administration were Wave 1 (March 10-30, 2022), Wave 2 (April 4-May 1, 2022), Wave 3 (May 4-June 2, 2022), Wave 4 (August 1-18, 2022), Wave 5 (September 26-October 15, 2022). Each wave consisted of approximately 5000 unique participants who were recruited to create samples that approximated population estimates for age, sex, race, and ethnicity based on the 2020 U.S. census. Surveys were conducted online by Qualtrics, LLC (Provo, Utah, and Seattle, Washington, U.S.), using their network of participant pools with varying recruitment methodologies. Informed consent was obtained electronically. The study was approved by the Monash University Human Research Ethics Committee (Study #24036).

### Survey Items

Participants self-reported demographic, anthropometric, and socioeconomic information including age, race, ethnicity, sex, height and weight, education level, employment status and household income. In addition, they provided information on several current and past medical conditions by answering the question: “Have you ever been diagnosed with any of the following conditions?” In addition to insomnia, opportunity was provided to endorse obstructive sleep apnea (OSA), high blood pressure, cardiovascular disease (e.g., heart attack, stroke, angina), gastrointestinal disorder (e.g., acid reflux, ulcers, indigestion), cancer, chronic kidney disease, liver disease, sickle cell disease, chronic obstructive pulmonary disease, and asthma. Possible responses to each condition were “Never”, “Yes I have in the past, but don’t have it now”, “Yes I have, but I do not regularly take medications or receiving treatment”, and “Yes I have, and I am regularly taking medications or receiving treatment”.

The following two questions pertaining to sleep quality were asked of the participants:

a. “Thinking about the past month, to what extent has poor sleep troubled you in general?”. Possible responses were “Not at all”, “A little”, “Somewhat”, “Much” and “Very Much”.
b. From the Pittsburgh Sleep Quality Index,^16^ “During the past month, how would you rate your sleep quality overall?” Possible responses were “Very good”, Fairly good”, “Fairly bad” and “Very bad”.

Sleep duration was assessed using a question from the Pittsburgh Sleep Quality Index.^16^ Responses were rounded to the nearest hour; those <3 hours or >12 hours were excluded as improbable estimates (N=987). In addition, sleep duration was stratified as short sleep (≤6 hours), recommended sleep (7-8 hours) and long sleep (≥9 hours).

Symptoms of OSA were obtained from responses to the Pittsburgh Sleep Quality Index and included items related to roommate or bedpartner reported “loud snoring” and “long pauses between breaths while you sleep”.^16^ In addition, sleepiness was assessed from the following item in the questionnaire: “During the past month, how often have you had trouble staying awake while driving, eating meals, or engaging in social activity”. Possible responses to all three items were “not during the past month”, “less than once a week”, once or twice a week” or “three or more times a week”. As done in our previous studies, participants were considered to have symptoms of OSA if they had either of the following combination of symptoms: 1) snoring “three or more times a week” and witnessed apnea or sleepiness “once or twice a week”; 2) witnessed apnea and sleepiness “once or twice a week”.^6,17^

Each survey contained identical items related to COVID-19 infection status and the number of COVID-19 vaccinations participants had received. Ascertainment of past COVID-19 infection was obtained using responses to the following questions related to COVID-19 testing or the presence of loss of taste or smell:

1. “Have you ever tested positive?”
2. “Despite never testing positive, are you confident that you have had COVID-19?”
3. “Despite never testing positive, have you received a clinical diagnosis of COVID-19?”
4. “Have you experienced a problem with decreased sense of smell or taste at any point since January 2020?”

COVID-19 vaccination status was ascertained by asking “How many COVID-19 vaccine doses have you received? (If you have had two doses of one brand and one of another, please select three)”. Participants were allowed to respond from 0 to 4.

### Statistical Analyses

<u>Exposures</u>: Based on our previous analyses in this cohort, we defined a positive history of COVID-19 infection as an affirmative response to having tested positive for COVID-19, a clinical diagnosis of COVID-19, or loss of taste or smell.^6,9,17^ Participants were classified as having OSA if they affirmed currently having the condition whether treated or not or if they had two or more symptoms of OSA.^6,9,17^ Insomnia was present if endorsed by participants irrespective of treatment status. Poor sleep quality was defined as being troubled by poor sleep “Much” or “Very Much” or rating their sleep quality as “Fairly Bad” or “Very Bad”.^9^

Vaccination status was dichotomized as Boosted (>2 vaccinations) or Not Boosted (≤2 vaccinations). Comorbid medical conditions were defined as currently having the condition whether treated or untreated. The effect of comorbid medical conditions was evaluated by summing the number of conditions reported by the participant (minimum value 0, maximum value 9).^6,9,17^ Body mass index (BMI) was calculated using self-reported height and weight as kg/m^2^. Socioeconomic covariates were dichotomized as follows: employment (retired vs. not retired), education (high school or less vs. some college) and income in U.S. Dollars (<$50,000 vs ≥$50,000).

Outcomes: Based on previous analyses of this cohort, PASC was defined as the presence of 3 or more symptoms commonly associated with PASC for at least 3 months after their COVID-19 infection (COPE).^6^ For comparison, 2 other definitions of PASC were also used: 1) a PASC Score ≥12 modified from the RECOVER cohort proposal,^13^ and 2) one or more symptoms commonly associated with PASC for at least 3 months after their COVID-19 infection as suggested by the United Kingdom’s National Institute for Health and Care Excellence (NICE).^12^ Because the COPE survey did not contain a few items comparable to those in the RECOVER cohort, we substituted symptoms contained in the COPE survey which were correlated^13^ or similar as follows: sore throat for thirst, sleep problems for sexual function, headache for dizziness, and other aches for abnormal movements.

<u>Analytics</u>: Summary data for continuous or ordinal variables are reported as their respective means and standard deviations (SD) and for categorical variables as their percentages. Comparisons of co-morbid medical, demographic, and social characteristic variables stratified by presence or absence of PASC were performed using Student’s unpaired t-test for continuous or ordinal variables and χ^2^ for categorical variables.

Multivariable modelling using logistic regression was utilized to determine whether insomnia, poor sleep quality and sleep duration were associated with the COPE, RECOVER and NICE models of PASC. For each definition, a baseline model was constructed using only insomnia, poor sleep quality and sleep duration stratified as short (≤6 hours), recommended (7-8 hours),^18^ and long (≥9 hours) sleep. We then developed increasingly complex models by sequentially including demographic factors, comorbidities, boosted vaccination status, socioeconomic factors, and other sleep conditions. Inasmuch as the sleep conditions evaluated in the models may have been correlated, diagnostic tests for collinearity were performed; the variance inflation factor was less than 10 and the tolerance was greater than 0.1 for all variables. However, because poor sleep quality can be considered a component of insomnia, models for insomnia were not adjusted for poor sleep quality. Conversely, models for poor sleep quality were not adjusted by insomnia. Results of the logistic regression models are presented as unadjusted or adjusted odds ratios (aOR) and their 95% confidence intervals (95% CI).

To determine whether our definition of COVID-19 infection status influenced our results, we performed sensitivity analyses with stricter (i.e., using COVID-19 infection as a positive test or loss of taste or smell) and broader (i.e., our original definition plus assumed positive for COVID-19 without a positive test or clinical diagnosis as an indicator of a past COVID-19 infection).

All analyses were conducted using IBM SPSS version 28 (Armonk, NY). A p<0.05 was considered statistically significant.

## Results

There were 24803 participants with evaluable data from the five COPE initiative surveys from which a positive history of COVID-19 infection (see Methods) was reported in 10324 (41.6%), which represented the analytic sample. Table 1 presents the demographic, anthropometric, co-morbid medical and social characteristics of these COVID-19 positive participants in the COPE cohort stratified by PASC status for the 3 models of PASC analyzed in this study. Characteristics of participants positive for PASC were generally similar for all 3 models. Those who were PASC positive were younger and more likely to be Hispanic, not retired, and have a higher income. They were less likely to have received a COVID-19 booster vaccination (COPE: 24.2% vs. 29.3%; NICE: 23.7% vs. 31.0%; PASC Score: 24.5% vs. 28.8%; all p<0.01). In terms of sleep, 3452 (45.6%) participants reported having slept the recommended 7-8 hours, 2973 (36.9%) screened positive for poor sleep quality, 1671 (20.7%) screened positive for insomnia, and 1697 (21.0%) screened positive for OSA. Compared with participants who did not experience PASC, those with PASC had a higher prevalence of OSA, insomnia, and poor sleep quality. In addition, they were more likely to report a short sleep duration whereas there was a slight tendency for those with PASC to report a long sleep duration.

**Table 1:**
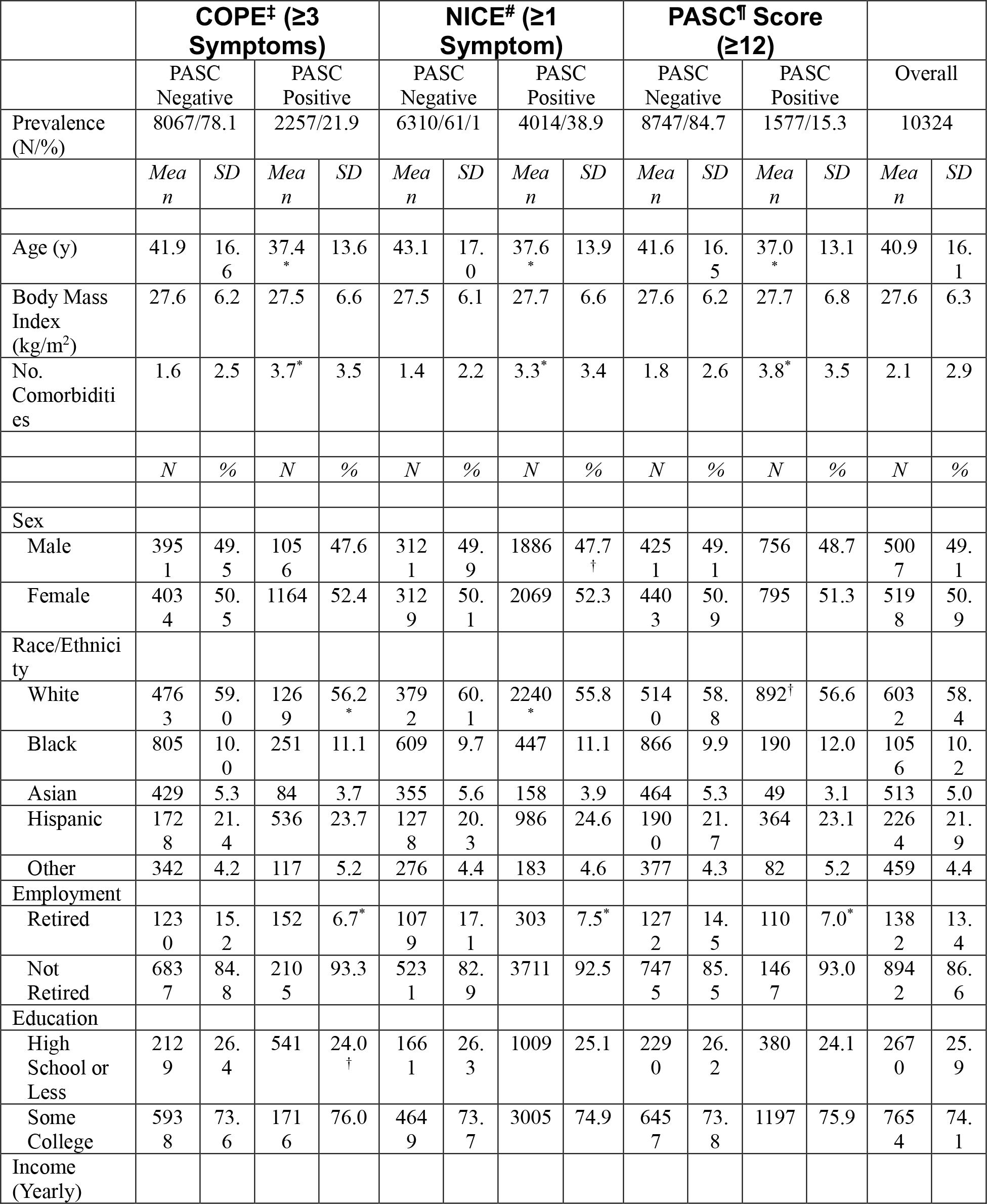
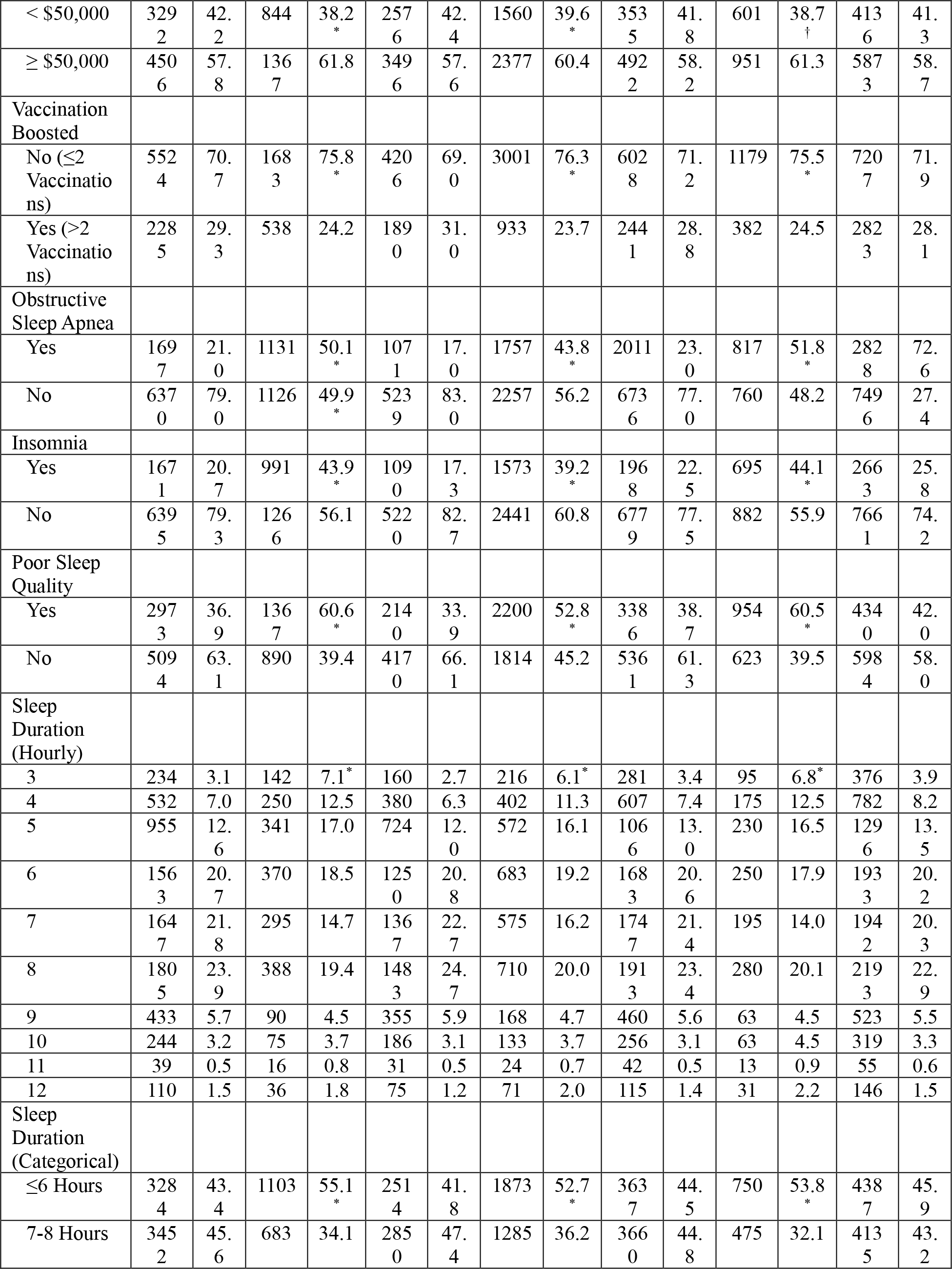
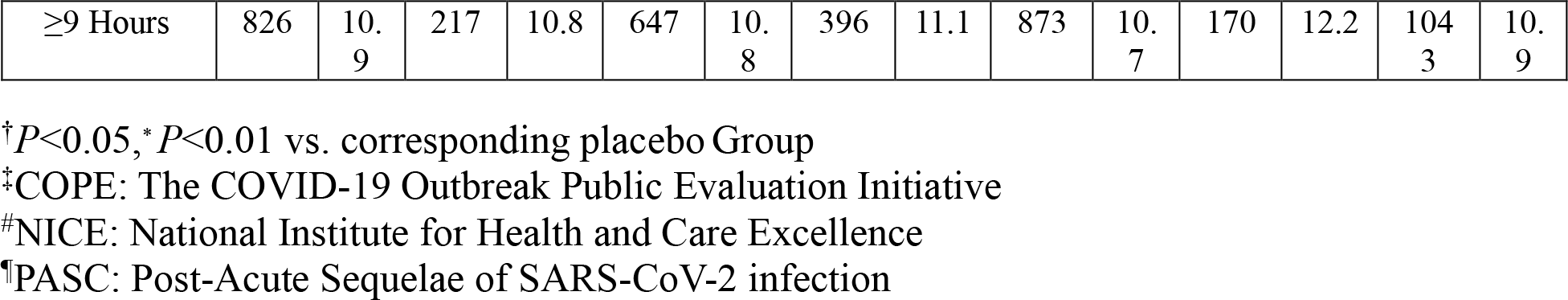
Associations Between PASC Status and Insomnia, Poor Sleep Quality, Sleep Duration, Co-morbid Medical, Demographic and Social Characteristics.

|  | COPE<sup>‡</sup> (≥3 Symptoms) |  |  |  | NICE<sup>#</sup> (≥1 Symptom) |  |  |  | PASC<sup>†</sup> Score (≥12) |  |  |  |  |  |
| --- | --- | --- | --- | --- | --- | --- | --- | --- | --- | --- | --- | --- | --- | --- |
|  | PASC Negative |  | PASC Positive |  | PASC Negative |  | PASC Positive |  | PASC Negative |  | PASC Positive |  | Overall |  |
| Prevalence (N/%) | 8067/78.1 |  | 2257/21.9 |  | 6310/61.1 |  | 4014/38.9 |  | 8747/84.7 |  | 1577/15.3 |  | 10324 |  |
|  | Mean | SD | Mean | SD | Mean | SD | Mean | SD | Mean | SD | Mean | SD | Mean | SD |
| Age (y) | 41.9 | 16.6 | 37.4* | 13.6 | 43.1 | 17.0 | 37.6* | 13.9 | 41.6 | 16.5 | 37.0* | 13.1 | 40.9 | 16.1 |
| Body Mass Index (kg/m<sup>2</sup>) | 27.6 | 6.2 | 27.5 | 6.6 | 27.5 | 6.1 | 27.7 | 6.6 | 27.6 | 6.2 | 27.7 | 6.8 | 27.6 | 6.3 |
| No. Comorbidities | 1.6 | 2.5 | 3.7* | 3.5 | 1.4 | 2.2 | 3.3* | 3.4 | 1.8 | 2.6 | 3.8* | 3.5 | 2.1 | 2.9 |
|  | N | % | N | % | N | % | N | % | N | % | N | % | N | % |
| Sex |  |  |  |  |  |  |  |  |  |  |  |  |  |  |
| Male | 3951 | 49.5 | 1056 | 47.6 | 3121 | 49.9 | 1886 | 47.7† | 4251 | 49.1 | 756 | 48.7 | 5007 | 49.1 |
| Female | 4034 | 50.5 | 1164 | 52.4 | 3129 | 50.1 | 2069 | 52.3 | 4403 | 50.9 | 795 | 51.3 | 5198 | 50.9 |
| Race/Ethnicity |  |  |  |  |  |  |  |  |  |  |  |  |  |  |
| White | 4763 | 59.0 | 1269 | 56.2* | 3792 | 60.1 | 2240* | 55.8 | 5140 | 58.8 | 892† | 56.6 | 6032 | 58.4 |
| Black | 805 | 10.0 | 251 | 11.1 | 609 | 9.7 | 447 | 11.1 | 866 | 9.9 | 190 | 12.0 | 1056 | 10.2 |
| Asian | 429 | 5.3 | 84 | 3.7 | 355 | 5.6 | 158 | 3.9 | 464 | 5.3 | 49 | 3.1 | 513 | 5.0 |
| Hispanic | 1728 | 21.4 | 536 | 23.7 | 1278 | 20.3 | 986 | 24.6 | 1900 | 21.7 | 364 | 23.1 | 2264 | 21.9 |
| Other | 342 | 4.2 | 117 | 5.2 | 276 | 4.4 | 183 | 4.6 | 377 | 4.3 | 82 | 5.2 | 459 | 4.4 |
| Employment |  |  |  |  |  |  |  |  |  |  |  |  |  |  |
| Retired | 1230 | 15.2 | 152 | 6.7* | 1079 | 17.1 | 303 | 7.5* | 1272 | 14.5 | 110 | 7.0* | 1382 | 13.4 |
| Not Retired | 6837 | 84.8 | 2105 | 93.3 | 5231 | 82.9 | 3711 | 92.5 | 7475 | 85.5 | 1467 | 93.0 | 8942 | 86.6 |
| Education |  |  |  |  |  |  |  |  |  |  |  |  |  |  |
| High School or Less | 2129 | 26.4 | 541 | 24.0† | 1661 | 26.3 | 1009 | 25.1 | 2290 | 26.2 | 380 | 24.1 | 2670 | 25.9 |
| Some College | 5938 | 73.6 | 1716 | 76.0 | 4649 | 73.7 | 3005 | 74.9 | 6457 | 73.8 | 1197 | 75.9 | 7654 | 74.1 |
| Income (Yearly) |  |  |  |  |  |  |  |  |  |  |  |  |  |  |
| < \$50,000 | 329<br>2 | 42.<br>2 | 844 | 38.2<br>* | 257<br>6 | 42.<br>4 | 1560 | 39.6<br>* | 353<br>5 | 41.<br>8 | 601 | 38.7<br>† | 413<br>6 | 41.<br>3 |
| ≥ \$50,000 | 450<br>6 | 57.<br>8 | 136<br>7 | 61.8 | 349<br>6 | 57.<br>6 | 2377 | 60.4 | 492<br>2 | 58.<br>2 | 951 | 61.3 | 587<br>3 | 58.<br>7 |
| Vaccination<br>Boosted |  |  |  |  |  |  |  |  |  |  |  |  |  |  |
| No (≤2<br>Vaccinatio<br>ns) | 552<br>4 | 70.<br>7 | 168<br>3 | 75.8<br>* | 420<br>6 | 69.<br>0 | 3001 | 76.3<br>* | 602<br>8 | 71.<br>2 | 1179 | 75.5<br>* | 720<br>7 | 71.<br>9 |
| Yes (>2<br>Vaccinatio<br>ns) | 228<br>5 | 29.<br>3 | 538 | 24.2 | 189<br>0 | 31.<br>0 | 933 | 23.7 | 244<br>1 | 28.<br>8 | 382 | 24.5 | 282<br>3 | 28.<br>1 |
| Obstructive<br>Sleep Apnea |  |  |  |  |  |  |  |  |  |  |  |  |  |  |
| Yes | 169<br>7 | 21.<br>0 | 1131 | 50.1<br>* | 107<br>1 | 17.<br>0 | 1757 | 43.8<br>* | 2011 | 23.<br>0 | 817 | 51.8<br>* | 282<br>8 | 72.<br>6 |
| No | 637<br>0 | 79.<br>0 | 1126 | 49.9<br>* | 523<br>9 | 83.<br>0 | 2257 | 56.2 | 673<br>6 | 77.<br>0 | 760 | 48.2 | 749<br>6 | 27.<br>4 |
| Insomnia |  |  |  |  |  |  |  |  |  |  |  |  |  |  |
| Yes | 167<br>1 | 20.<br>7 | 991 | 43.9<br>* | 109<br>0 | 17.<br>3 | 1573 | 39.2<br>* | 196<br>8 | 22.<br>5 | 695 | 44.1<br>* | 266<br>3 | 25.<br>8 |
| No | 639<br>5 | 79.<br>3 | 126<br>6 | 56.1 | 522<br>0 | 82.<br>7 | 2441 | 60.8 | 677<br>9 | 77.<br>5 | 882 | 55.9 | 766<br>1 | 74.<br>2 |
| Poor Sleep<br>Quality |  |  |  |  |  |  |  |  |  |  |  |  |  |  |
| Yes | 297<br>3 | 36.<br>9 | 136<br>7 | 60.6<br>* | 214<br>0 | 33.<br>9 | 2200 | 52.8<br>* | 338<br>6 | 38.<br>7 | 954 | 60.5<br>* | 434<br>0 | 42.<br>0 |
| No | 509<br>4 | 63.<br>1 | 890 | 39.4 | 417<br>0 | 66.<br>1 | 1814 | 45.2 | 536<br>1 | 61.<br>3 | 623 | 39.5 | 598<br>4 | 58.<br>0 |
| Sleep<br>Duration<br>(Hourly) |  |  |  |  |  |  |  |  |  |  |  |  |  |  |
| 3 | 234 | 3.1 | 142 | 7.1* | 160 | 2.7 | 216 | 6.1* | 281 | 3.4 | 95 | 6.8* | 376 | 3.9 |
| 4 | 532 | 7.0 | 250 | 12.5 | 380 | 6.3 | 402 | 11.3 | 607 | 7.4 | 175 | 12.5 | 782 | 8.2 |
| 5 | 955 | 12.<br>6 | 341 | 17.0 | 724 | 12.<br>0 | 572 | 16.1 | 106<br>6 | 13.<br>0 | 230 | 16.5 | 129<br>6 | 13.<br>5 |
| 6 | 156<br>3 | 20.<br>7 | 370 | 18.5 | 125<br>0 | 20.<br>8 | 683 | 19.2 | 168<br>3 | 20.<br>6 | 250 | 17.9 | 193<br>3 | 20.<br>2 |
| 7 | 164<br>7 | 21.<br>8 | 295 | 14.7 | 136<br>7 | 22.<br>7 | 575 | 16.2 | 174<br>7 | 21.<br>4 | 195 | 14.0 | 194<br>2 | 20.<br>3 |
| 8 | 180<br>5 | 23.<br>9 | 388 | 19.4 | 148<br>3 | 24.<br>7 | 710 | 20.0 | 191<br>3 | 23.<br>4 | 280 | 20.1 | 219<br>3 | 22.<br>9 |
| 9 | 433 | 5.7 | 90 | 4.5 | 355 | 5.9 | 168 | 4.7 | 460 | 5.6 | 63 | 4.5 | 523 | 5.5 |
| 10 | 244 | 3.2 | 75 | 3.7 | 186 | 3.1 | 133 | 3.7 | 256 | 3.1 | 63 | 4.5 | 319 | 3.3 |
| 11 | 39 | 0.5 | 16 | 0.8 | 31 | 0.5 | 24 | 0.7 | 42 | 0.5 | 13 | 0.9 | 55 | 0.6 |
| 12 | 110 | 1.5 | 36 | 1.8 | 75 | 1.2 | 71 | 2.0 | 115 | 1.4 | 31 | 2.2 | 146 | 1.5 |
| Sleep<br>Duration<br>(Categorical) |  |  |  |  |  |  |  |  |  |  |  |  |  |  |
| ≤6 Hours | 328<br>4 | 43.<br>4 | 1103 | 55.1<br>* | 251<br>4 | 41.<br>8 | 1873 | 52.7<br>* | 363<br>7 | 44.<br>5 | 750 | 53.8<br>* | 438<br>7 | 45.<br>9 |
| 7-8 Hours | 345<br>2 | 45.<br>6 | 683 | 34.1 | 285<br>0 | 47.<br>4 | 1285 | 36.2 | 366<br>0 | 44.<br>8 | 475 | 32.1 | 413<br>5 | 43.<br>2 |
| ≥9 Hours | 826 | 10.<br>9 | 217 | 10.8 | 647 | 10.<br>8 | 396 | 11.1 | 873 | 10.<br>7 | 170 | 12.2 | 104<br>3 | 10.<br>9 |

<sup>†</sup> $P<0.05$ , \* $P<0.01$ vs. corresponding placebo Group
<sup>‡</sup>COPE: The COVID-19 Outbreak Public Evaluation Initiative
<sup>#</sup>NICE: National Institute for Health and Care Excellence
<sup>¶</sup>PASC: Post-Acute Sequelae of SARS-CoV-2 infection

The prevalence of PASC as defined in the COPE, NICE, and PASC score models is shown in Table 1. The most restrictive model was the PASC score (15.3%) followed by the COPE (21.9%) and NICE (38.9%) models.

In Table 2 are logistic regression analyses describing among participants with a positive history of COVID-19 infection the association between insomnia and PASC in all 3 PASC models at baseline and subsequently in partially and fully adjusted models. The aORs in the fully adjusted models were comparable with narrow confidence intervals (CI) ranging from 1.30 (95% CI: 1.11-1.52, p≤0.05, PASC Score) to 1.52 (95% CI: 1.34-1.71, p≤0.001, (NICE). In none of the models did boosted vaccination affect the relationship between insomnia and PASC (insomnia x boosted vaccination aORs were not significant, data not shown).

**Table 2:**
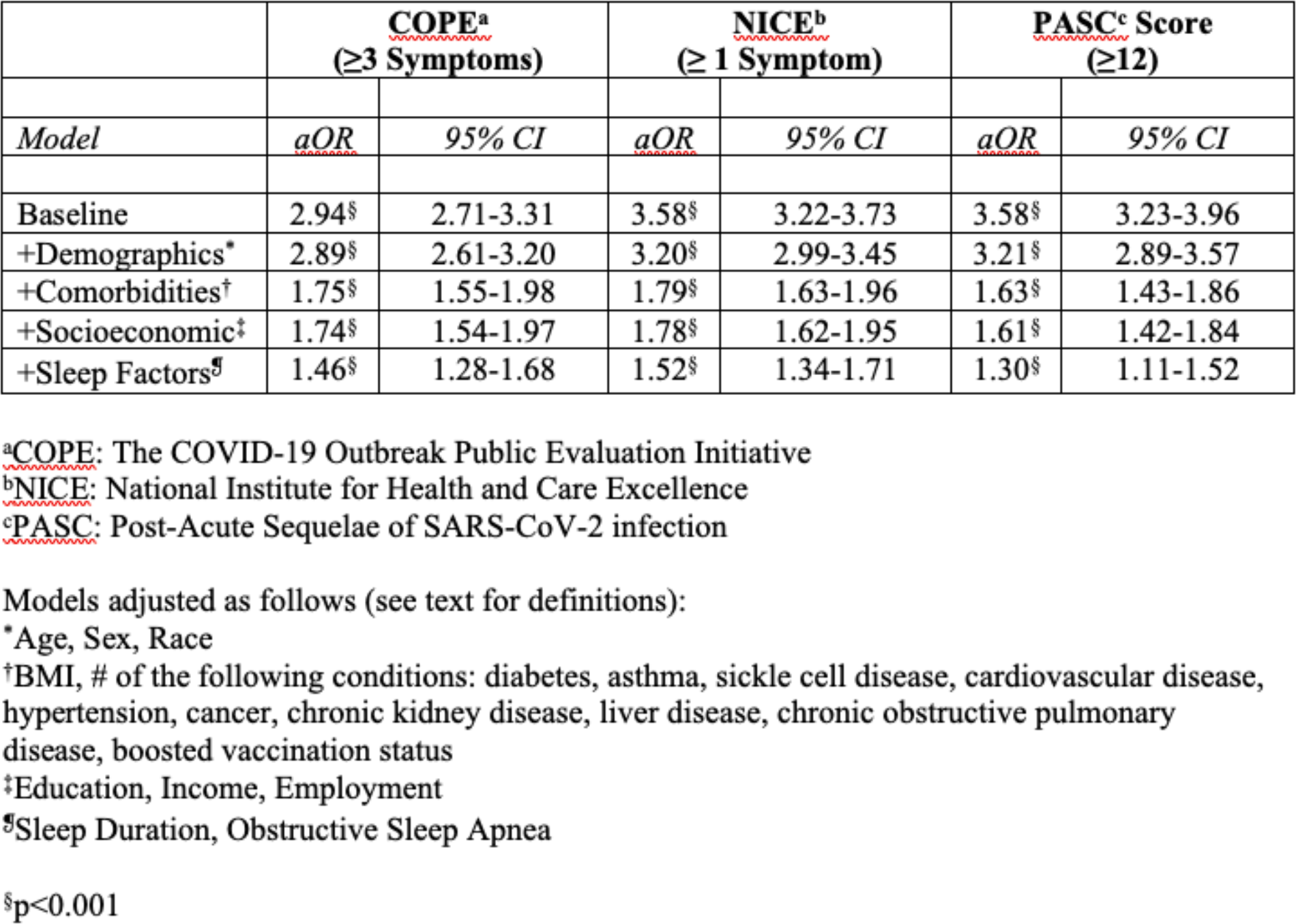
Association of Self-reported Insomnia with Three Models of PASC.

|  | **COPE<sup>a</sup>**<br>(≥3 Symptoms) |  | **NICE<sup>b</sup>**<br>(≥ 1 Symptom) |  | **PASC<sup>c</sup> Score**<br>(≥12) |  |
| --- | --- | --- | --- | --- | --- | --- |
| *Model* | *aOR* | *95% CI* | *aOR* | *95% CI* | *aOR* | *95% CI* |
| Baseline | 2.94<sup>§</sup> | 2.71-3.31 | 3.58<sup>§</sup> | 3.22-3.73 | 3.58<sup>§</sup> | 3.23-3.96 |
| +Demographics* | 2.89<sup>§</sup> | 2.61-3.20 | 3.20<sup>§</sup> | 2.99-3.45 | 3.21<sup>§</sup> | 2.89-3.57 |
| +Comorbidities<sup>†</sup> | 1.75<sup>§</sup> | 1.55-1.98 | 1.79<sup>§</sup> | 1.63-1.96 | 1.63<sup>§</sup> | 1.43-1.86 |
| +Socioeconomic<sup>‡</sup> | 1.74<sup>§</sup> | 1.54-1.97 | 1.78<sup>§</sup> | 1.62-1.95 | 1.61<sup>§</sup> | 1.42-1.84 |
| +Sleep Factors<sup>§</sup> | 1.46<sup>§</sup> | 1.28-1.68 | 1.52<sup>§</sup> | 1.34-1.71 | 1.30<sup>§</sup> | 1.11-1.52 |

<sup>a</sup>COPE: The COVID-19 Outbreak Public Evaluation Initiative<sup>b</sup>NICE: National Institute for Health and Care Excellence<sup>c</sup>PASC: Post-Acute Sequelae of SARS-CoV-2 infection
Models adjusted as follows (see text for definitions):
\*Age, Sex, Race
<sup>†</sup>BMI, # of the following conditions: diabetes, asthma, sickle cell disease, cardiovascular disease, hypertension, cancer, chronic kidney disease, liver disease, chronic obstructive pulmonary disease, boosted vaccination status<sup>‡</sup>Education, Income, Employment<sup>§</sup>Sleep Duration, Obstructive Sleep Apnea<sup>§</sup>p<0.001

Table 3 describes the association of poor sleep quality with PASC among participants with a positive history of COVID-19 infection. For all 3 models, poor sleep quality was related to an increased probability of PASC; aORs at baseline were comparable ranging from 2.36 (95% CI: 2.18-2.56, p≤0.001, NICE) to 2.63 (95% CI: 2.39-2.90, p≤0.001, COPE). Subsequent adjustment for demographic and socioeconomic factors, and sleep conditions as well as comorbidities attenuated these associations, but all remained significant in fully adjusted models with aORs ranging from 1.77 (95% CI: 1.60-1.97, p≤0.001, NICE) to 2.00 (95% CI: 1.77-2.26, p≤0.001, COPE). In none of the models did boosted vaccination affect the relationship between poor sleep quality and PASC (poor sleep quality x boosted vaccination aORs were not significant, data not shown).

**Table 3:**
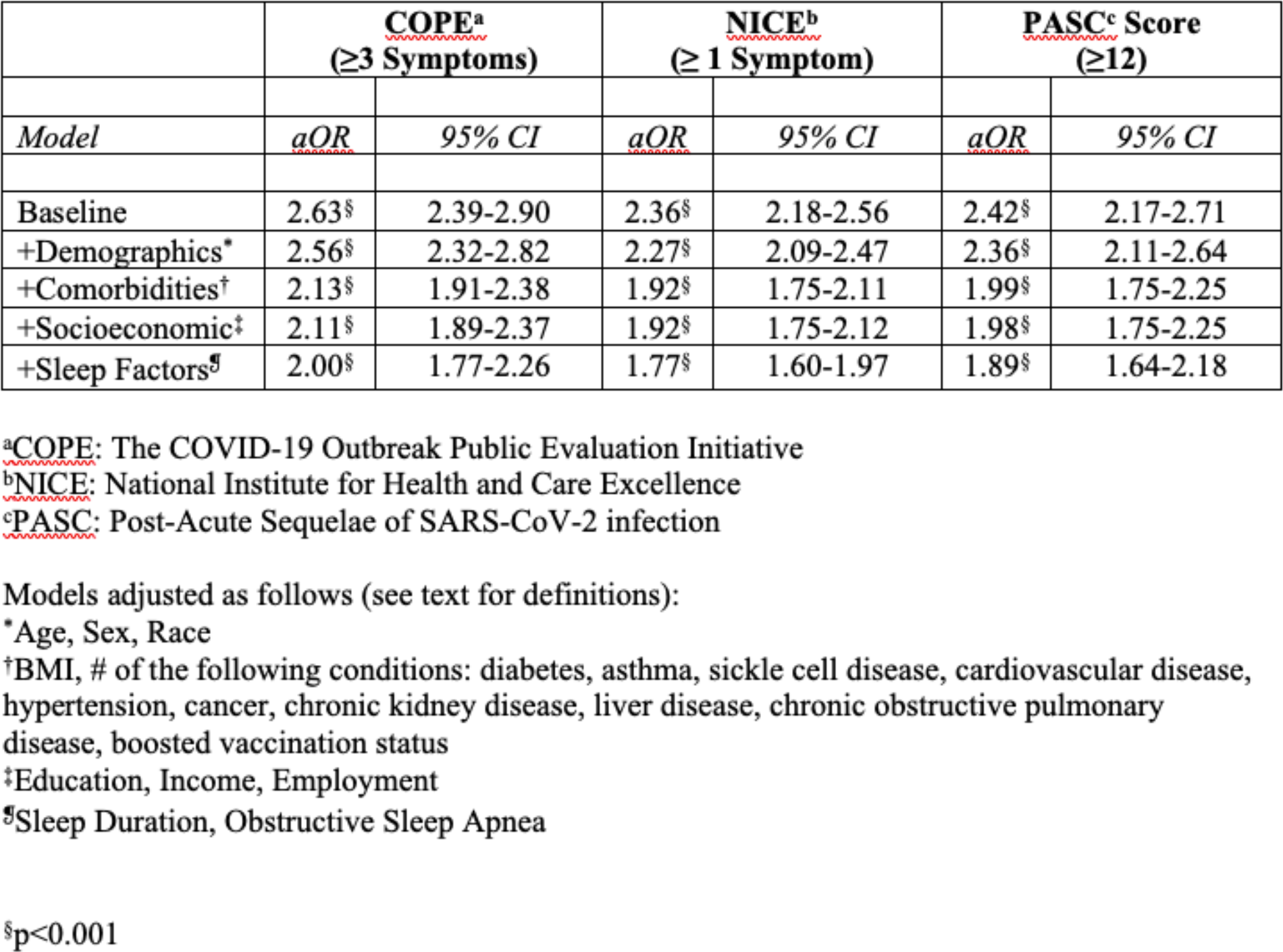
Association of Poor Sleep Quality with Three Models of PASC.

In Table 4 are shown the relationships between the full distribution of hourly sleep duration (3 to 12 hours) with 3 models of PASC among participants with a positive history of COVID-19 infection. For all 3 models, the unadjusted aORs suggested that both short and long sleep durations were associated with PASC in comparison to the recommended 7-8 hours of sleep. For sleep durations less than 7-8 hours, these findings were attenuated, but remained significant. However, for sleep durations greater than 7-8 hours, the association with PASC was no longer present with the solitary exception of 11 hours in the PASC Score model.

**Table 4:**
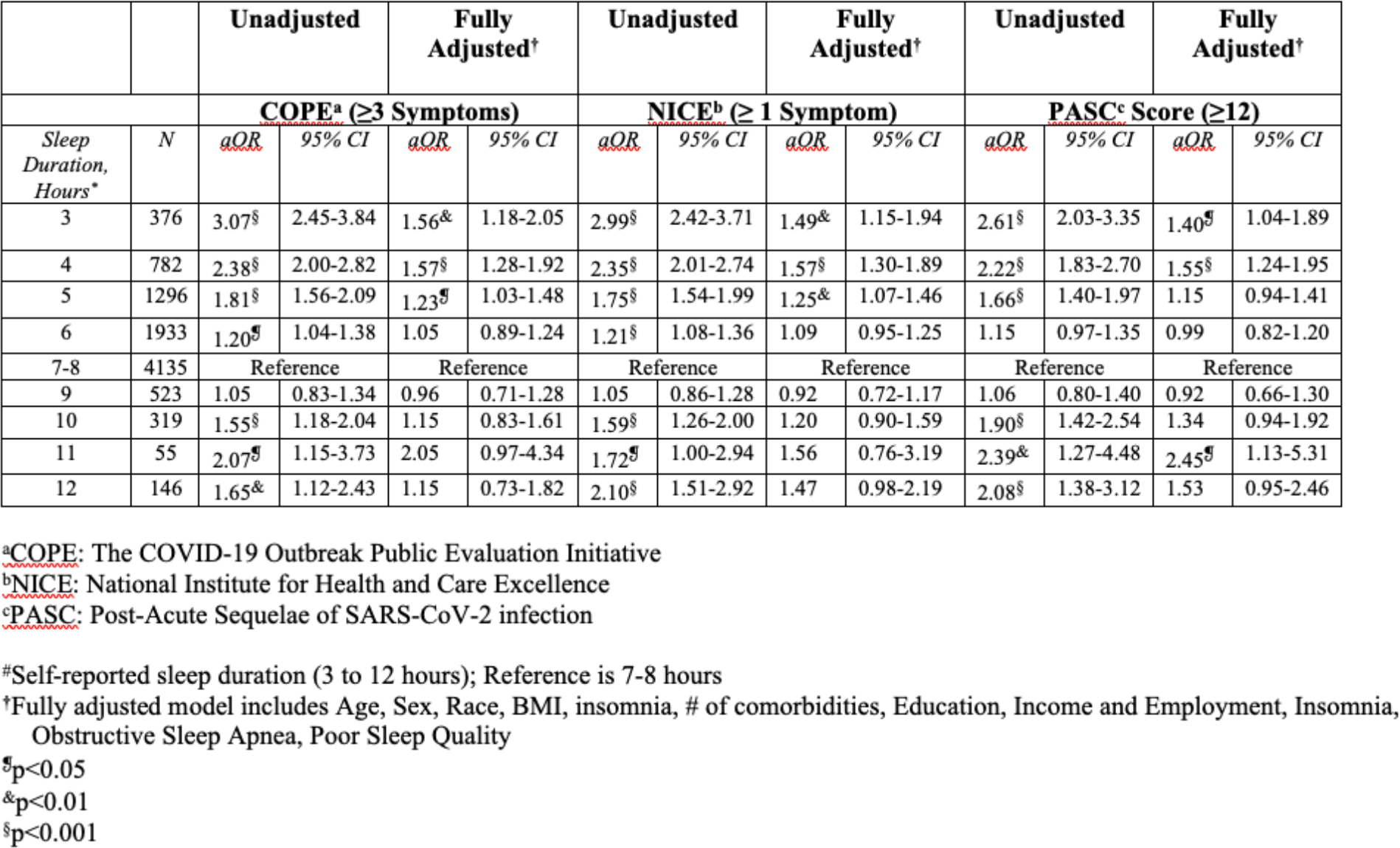
Odds ratio (adjusted) for association of self-reported hourly sleep duration* with 3 models of PASC.

Shown in Table 5 are the associations of short sleep (≤6 hours) and long sleep (≥9 hours) with PASC among participants with a positive history of COVID-19 infection. In comparison to recommended hours of sleep (7-8 hours; n = 3452), short sleep (n = 3284) was associated with a greater likelihood of PASC. The aORs at baseline were comparable for all 3 models and ranged from 1.59 (95% CI: 1.40-1.80, p≤0.001, PASC Score) to 1.70 (95% CI: 1.53-1.89, p≤0.001, COPE). These associations were attenuated but remained significant in fully adjusted models with aORs of 1.17 (95% CI: 1.01-1.36, p≤0.05, PASC Score), 1.23 (95% CI: 1.08-1.41, p≤0.001, COPE), and 1.23 (95% CI: 1.10-1.37, p≤0.001, NICE). Long sleep (n = 826) also was associated with a higher probability of PASC at baseline in all 3 models with aORs ranging from 1.33 (95% CI: 1.12-1.58, p≤0.001, COPE) to 1.50 (95% CI: 1.24-1.82, p≤0.001, PASC Score). However, in contrast to short sleep, adjustment for demographic and socioeconomic factors, and sleep conditions as well as comorbidities negated these associations in fully adjusted models. The associations between short and long sleep, and all 3 models of PASC were not mitigated by having received a COVID-19 booster (sleep duration x boosted vaccination aORs were not significant, data not shown).

**Table 5:** Association of Self-reported Sleep Duration with Three Models of PASC.

|  | **COPE<sup>a</sup>**<br>**(≥3 Symptoms)** |  | **NICE<sup>b</sup>**<br>**(≥ 1 Symptom)** |  | **PASC Score<sup>c</sup>**<br>**(≥12)** |  |
| --- | --- | --- | --- | --- | --- | --- |
| *Model* | *aOR* | *95% CI* | *aOR* | *95% CI* | *aOR* | *95% CI* |
| **SLEEP ≤6 HOURS<sup>#</sup>** |  |  |  |  |  |  |
| Unadjusted | 1.70<sup>§</sup> | 1.53-1.89 | 1.62<sup>§</sup> | 1.51-1.81 | 1.59<sup>§</sup> | 1.40-1.80 |
| +Demographics<sup>*</sup> | 1.66<sup>§</sup> | 1.49-1.85 | 1.61<sup>§</sup> | 1.47-1.76 | 1.56<sup>§</sup> | 1.37-1.76 |
| +Comorbidities<sup>†</sup> | 1.52<sup>§</sup> | 1.35-1.72 | 1.48<sup>§</sup> | 1.33-1.64 | 1.41<sup>§</sup> | 1.23-1.63 |
| +Socioeconomic<sup>‡</sup> | 1.55<sup>§</sup> | 1.37-1.75 | 1.51<sup>§</sup> | 1.36-1.68 | 1.43<sup>§</sup> | 1.24-1.65 |
| +Sleep Factors<sup>§</sup> | 1.23<sup>§</sup> | 1.08-1.41 | 1.23<sup>§</sup> | 1.10-1.37 | 1.17<sup>§</sup> | 1.01-1.36 |
| **SLEEP ≥9 HOURS<sup>#</sup>** |  |  |  |  |  |  |
| Unadjusted | 1.33<sup>§</sup> | 1.12-1.58 | 1.36<sup>§</sup> | 1.18-1.56 | 1.50<sup>§</sup> | 1.24-1.82 |
| +Demographics<sup>¶</sup> | 1.20<sup>¶</sup> | 1.01-1.43 | 1.22<sup>&</sup> | 1.05-1.41 | 1.35<sup>&</sup> | 1.11-1.64 |
| +Comorbidities<sup>†</sup> | 1.05 | 0.86-1.28 | 1.05 | 0.89-1.24 | 1.18 | 0.95-1.48 |
| +Socioeconomic<sup>‡</sup> | 1.12 | 0.92-1.38 | 1.11 | 0.94-1.32 | 1.25 | 0.99-1.56 |
| +Sleep Factors<sup>§</sup> | 1.12 | 0.91-1.37 | 1.12 | 0.94-1.33 | 1.24 | 0.99-1.55 |

<sup>a</sup>COPE: The COVID-19 Outbreak Public Evaluation Initiative<sup>b</sup>NICE: National Institute for Health and Care Excellence<sup>c</sup>PASC: Post-Acute Sequelae of SARS-CoV-2 infection
Models adjusted as follows (see text for definitions):
<sup>#</sup>Referent is 7-8 Hours<sup>\*</sup>Age, Sex, Race<sup>†</sup>BMI, # of the following conditions: diabetes, asthma, sickle cell disease, cardiovascular disease, hypertension, cancer, chronic kidney disease, liver disease, chronic obstructive pulmonary disease, boosted vaccination status<sup>‡</sup>Education, Income, Employment<sup>§</sup>Insomnia, Obstructive Sleep Apnea, Sleep Quality<sup>¶</sup>p≤0.05; <sup>&</sup>p≤0.01, <sup>§</sup>p<0.001

In sensitivity analyses, using a stricter definition of COVID-19 that required a positive test or loss of taste or smell, the positive associations between insomnia, poor sleep quality and short sleep duration remained (data not shown). Similarly, using a less stringent definition of COVID-19 that allowed for a positive diagnosis for any unconfirmed report of infection did not alter these relationships (data not shown).

## Discussion

In this study, the prevalence of PASC among participants with a positive history of COVID-19 infection was estimated in a large U.S. general population cohort using three different models of PASC and found to range between 15.3% and 38.9%. In all three models, insomnia, poor sleep quality and sleep shorter than recommended levels were associated with PASC after adjustment for demographic, anthropometric, and socioeconomic factors, and comorbid medical conditions. In contrast, sleep longer than recommended was not related to PASC.

Although it is generally acknowledged that a substantial proportion of persons who develop acute COVID-19 infection have a multitude of lingering symptoms, there is no agreement on a unifying definition of PASC. Such a definition is important for public health case identification as well as for guiding future research. However, several definitions have been proposed using different requirements for number of symptoms and time elapsed after acute infection. In the current study, we estimated the prevalence of PASC using a definition proposed by NICE in the United Kingdom^12^ and another modified from the RECOVER cohort in the U.S.^13^ and compared them to one we developed for the COPE Initiative.^6^ We found that among participants with a positive history of COVID-19 infection, prevalence estimates of PASC were 21.9%, 38.9% and 15.3% using the COPE, NICE, and RECOVER models, respectively. In contrast, prevalence rates of 21.21% and 8.68% were observed using the NICE and PASC Score definitions in a survey of the adult population of Mexico.^19^ Analysis of the Behavioral Risk Factor Surveillance System (BRFSS) found a 21.7% prevalence using the NICE definition.^20^ The prevalence of PASC in the RECOVER cohort was 23%,^13^ but a much higher rate of 61% was noted in a study from the International COVID Sleep Study II (ICOSSII) group using the World Health Organization definition of at least one symptom lasting at least 3 months.^21^ Notably, a meta-analysis of a heterogeneous array of 33 studies found an overall prevalence of 37% 30 days after infection with a 95% confidence interval between 26 and 49%.^22^ The explanation for the considerable heterogeneity in PASC prevalence rates is not entirely clear, but investigations differed in both methods used to ascertain COVID-19 positivity and definition of PASC as well as in the population evaluated. Most studies utilized self-report of both COVID-19 infection and PASC associated symptoms,^6,20,21,23,24^ but in some, objective documentation of infection was required resulting in lower prevalence rates.^13,19^ To our knowledge, this is the first study in the U.S. to estimate prevalence rates of PASC using different definitions in the same cohort. However, the most appropriate definition to ultimately use may depend on which one best predicts clinical outcomes or responses to therapy.

We observed that among participants with a positive history of COVID-19 infection, both insomnia and poor sleep quality were strongly associated with PASC in all three of the models evaluated. With respect to insomnia, our results are consistent with data from a longitudinal study of 2,759 Italians in which higher pre-COVID-19 insomnia severity index scores were found to predict greater risk of individual symptoms of PASC 1 to 3 months after COVID-19 infection.^25^ Our findings extend this previous report by demonstrating in a larger cohort that insomnia is associated with several global definitions of PASC. Additionally, our results related to poor sleep quality are supported by data from the aforementioned Italian cohort study which showed that higher scores on the Pittsburgh Sleep Quality Index predicted a greater risk for developing symptoms associated with PASC.^25^ Furthermore, in a study of 1,581 persons from the United Kingdom who were surveyed concerning their sleep quality one month before their COVID-19 infection, persons who reported average to very poor sleep quality were 2.4-3.5 times more likely to self-report having “Long COVID”.^24^ Our study extends these previous observations by using a more precise definition of poor sleep quality as well as a more formal definitions of PASC.

Sleep duration ≤6 hours per night in comparison to recommended levels of 7-8 hours was found to be associated with PASC in all 3 models. Our findings replicate those noted in analyses of the BRFSS dataset in which <7 hours of sleep were also associated with higher odds of PASC.^20^ However, they differ from a report from the the International COVID Sleep Study II (ICOSSII) group who noted increased likelihood of PASC in those sleeping <6 hours per night only among individuals who had pre-existing medical co-morbidities; no association was observed in the absence of medical co-morbidities.^21^ In contrast to the strong association between short sleep duration and PASC, no such linkage was observed in our study for long sleep duration. Thus, our results with respect to long sleep duration are consistent with reports from the BRFSS and ICOSSII studies which also did not find increased odds of PASC with longer than recommended sleep duration.^20,21^

We observed that for all 3 PASC models, the prevalence of having received a COVID-19 booster vaccination was lower in persons with PASC. This finding is consistent with a meta-analysis of 18 studies ^26^ as well as with a recent large retrospective matched cohort study demonstrating that COVID-19 vaccination decreased the risk of developing PASC.^27^ Previous results from the COPE initiative indicated that receiving a COVID-19 booster vaccination reduced the risk of COVID-19 hospitalization attributable to OSA.^17^ In contrast, we did not find that boosted vaccination for COVID-19 affected the odds of developing PASC related to insomnia, poor sleep quality or short sleep duration.

There are several mechanisms that could explain the association between insomnia, poor sleep quality and short sleep duration, and PASC. Insomnia and most likely poor sleep quality are characterized by a state of stress and hyperarousal.^28^ This can lead to alterations in the hypothalamic-pituitary-adrenal axis related to inflammation; in a meta-analysis, sleep disturbance was associated with higher levels of C Reactive Protein and Interleukin-6.^29^ Reduced sleep can result in an increase in transcriptional pathways and greater cellular production of inflammatory cytokines from monocytes.^30^ Furthermore, it has been suggested that inflammatory processes are important in the pathogenesis of the dysautonomia, endothelial dysregulation, and neurocognitive impairments observed with PASC.^31^ Sleep disturbances also have been implicated in dysfunction of the immune system. Chronic sleep loss and disturbed sleep promote not only a proinflammatory state, but also affect the number and function of immune regulating cells ^32,33^ and prolong shedding of SARS-CoV-2 after infection.^34^ Both factors may promote viral persistence in tissues which is hypothesized as a mechanism resulting in PASC.^31^

Our findings that sleep disruption is associated with a greater likelihood of PASC suggest interventions improving sleep may mitigate or reduce the impact of PASC on overall public health. For example, information pertaining to the benefits of healthy sleep could be disseminated to persons who develop COVID-19 infection. Inasmuch as the prevalence of PASC in our study as well as others ranges between 26% and 49%,^22^ even a small reduction in the incidence of PASC could have profound public health importance.

There are several limitations to the interpretation of our findings. First, sleep disturbances can occur *de novo* after acute COVID-19 infection.^35,36^ Our analyses are cross-sectional, and therefore a conclusion that insomnia, poor sleep quality and short sleep duration are risk factors for the development of PASC should be interpreted cautiously. Second, sleep duration was self-reported. Self-reported sleep has historically overestimated sleep duration compared with the gold-standard objective measurement by polysomnography.^37^ However, inasmuch as both 3 and 4 hours of sleep in this study were associated with PASC, it is unlikely that any overestimates of sleep duration would have been sufficiently extreme to alter our findings of an association between short sleep duration and PASC. Third, the sample could be susceptible to misclassification bias with respect to history of COVID-19 infection, as infection with COVID-19 was based on self-report and included dyssomnia and dysgeusia. However, in sensitivity analyses our findings were robust when incorporating less and more stringent criteria. Finally, the COPE survey did not include all of the symptoms included in the RECOVER PASC score, which had not been published at the time the COPE survey was developed and initially administered. Nevertheless, the findings observed with our modification of the PASC Score were consistent with the COPE and NICE definitions suggesting that they were similarly representative of participants with PASC.

In summary, the prevalence of PASC ranged between 15.3% and 38.9% in persons who previously had a COVID-19 infection. Insomnia, poor sleep quality, and short sleep duration are associated with higher odds of PASC. Boosted vaccination for COVID-19 did not affect these observations. These findings emphasize the importance of sleep disturbance in COVID-19 infection and PASC and suggest that interventions to improve sleep may reduce the incidence of PASC.

## Author Approval

All authors have seen and approved the manuscript.

## Preprint Information

A preprint of this manuscript has posted on medRxiv:

## Acknowledgments

Concept and Design: SFQ

Data collection: MDW, MÉC, MEH

Data analysis and interpretation: SFQ, MEH, CFM, MLJ, MEH

Drafting of the manuscript: SFQ

Critical feedback and revision of manuscript: SFQ, MDW, MÉC, LKB, LAB, MEH, MLJ, RL, CFM, AR, RR, PV, SMWR, CAC

## Data Availability

All data produced in the present study are available upon reasonable request to the authors.

